# Infant gonadotropins predict spontaneous puberty in girls with Turner syndrome

**DOI:** 10.1101/2024.10.22.24315884

**Authors:** Alexandra Sawyer, Samantha Bothwell, Karli Swenson, Sharon Travers, Shanlee Davis

## Abstract

**Introduction:** Hypergonadotropic hypogonadism is a characteristic clinical manifestation of Turner syndrome (TS). While up to 30% and 20% of people with TS will have spontaneous thelarche and menarche respectively, there is a lack of evidence to predict who will retain sufficient ovarian function to achieve these outcomes. The aim of this study was to determine if follicle-stimulation hormone (FSH) and/or luteinizing hormone (LH) concentrations measured in infancy would accurately predict later spontaneous thelarche or menarche.

**Methods:** Patients with a diagnosis of Turner syndrome with FSH and/or LH clinically measured prior to three years of age and now ≥10 years of age with documented pubertal assessment were included (n=33). Differences in infant gonadotropin values were determined for patients with vs without spontaneous thelarche/menarche using Kruskal-Wallis tests. The optimal threshold of infant LH and FSH to predict spontaneous thelarche and menarche was then determined by maximizing the sum of sensitivity and specificity.

**Results:** The prevalence of spontaneous thelarche and menarche were 21.2% and 15.2% respectively. An infant LH value greater than 0.5 mIU/mL predicted lack of spontaneous thelarche with an estimated accuracy of 94% and lack of spontaneous menarche with an estimated accuracy of 96%. An infant FSH value greater than 37.4 mIU/mL predicted lack of lack of spontaneous thelarche with an accuracy of 97% and lack of spontaneous menarche with an accuracy of 100%.

**Conclusion:** Infant gonadotropin concentrations accurately predict spontaneous later thelarche and menarche for persons with TS.

## Introduction

Turner Syndrome (TS) is the partial or complete absence of the second sex chromosome in a phenotypic female associated with at least one or more of its characteristic clinical manifestations.(1) TS occurs in ∼1 in every 2,000 births, and although the average age of diagnosis is in late childhood, ascertainment in infancy is becoming much more common with the global adoption of prenatal cell free DNA screening.(2) One of the most common clinical manifestations is primary ovarian insufficiency (POI) leading to hypergonadotropic hypogonadism and infertility. Most females with TS will have POI prior to pubertal onset and will require estradiol to induce puberty, however some girls will have sufficient ovarian function remaining to start or even complete puberty.(1,3,4) Studies have estimated spontaneous thelarche (ST) in TS to be as high as 32% and spontaneous menarche (SM) around 20%, with the lowest rates in girls with non-mosaic monosomy X karyotype.(5) While spontaneous pregnancies in individuals with TS are rare, there is potential for fertility preservation through oocyte stimulation and cryopreservation or even ovarian tissue cryopreservation, although the appropriate candidates and timing for these procedures are currently unknown.(6)

Non-mosaic 45,X karyotype is the best predictor for the absence of spontaneous puberty and fertility in TS, however this does not allow individualization for counseling and early management of POI. Gonadotropins, including follicle stimulating hormone (FSH) and luteinizing hormone (LH), are secreted by the anterior pituitary gland in response to gonadotropin releasing hormone pulsatility in infancy and again at pubertal onset.(7) This biphasic pattern of LH and FSH has been shown in girls with TS as well, often with elevated values consistent with hypergonadotropic hypogonadism.(8) However, the lack of longitudinal data from an adequate number of girls makes the utility of infant gonadotropin concentrations unclear. It is not routinely recommended to measure gonadotropins in infancy, but these data could offer an opportunity for individualization in clinical care if they accurately predicted ovarian function later in life.(1)

Serum anti-Mullerian hormone (AMH) is another biomarker that can potentially be measured in TS to assess ovarian function as it is made by the primordial follicles in the ovary.(9-11) In healthy individuals, AMH has been shown to be relatively stable over childhood, with early childhood AMH correlating with the number of follicles at the time of puberty, as well inversely with FSH.(10,12) In TS specifically, a cross-sectional study has shown that a measurable serum AMH is significantly associated with an increased odds of spontaneous thelarche and menarche.(10) Furthermore, detectable AMH is associated with viable follicles on ovarian tissue samples, therefore this value is currently clinically important for fertility counseling and timing of fertility preservation.(13) Whether infant gonadotropin concentrations predict future childhood AMH has not yet been investigated, however would be useful for fertility counseling.

The objective of this study was to evaluate the utility of infant gonadotropins in predicting longitudinal ovarian function in girls with TS. We hypothesized that elevated FSH and LH in infancy would be predictive of later premature ovarian insufficiency as measured by lack of spontaneous thelarche and menarche and undetectable childhood AMH.

## Methods

Individual medical record review was conducted for all patients with TS seen at Children’s Hospital Colorado between 2009 and 2023 with FSH and/or LH results measured prior to 3 years of age and pubertal assessment ≥10 years of age. Patients with a history of gonadectomy were excluded. Clinically obtained serum gonadotropins were measured by chemiluminescent immunoassay and AMH by enzyme-linked immunosorbent assay in CLIA-approved laboratories. The first LH and/or FSH value prior to age 3 years was used for infant gonadotropins; the first values ≥10 years was used for the pubertal-age gonadotropin levels. For dichotomized analyses, gonadotropin concentrations were defined as elevated if they were above the laboratory cut-off values. We used the first childhood AMH value obtained >3 years of age and dichotomized these results to either detectable or undetectable. Spontaneous thelarche was defined as breast development documented by a clinician prior to initiation of any hormone replacement therapy, and spontaneous menarche was defined as vaginal bleeding prior to any exogenous hormone exposure.

The aim of the primary analysis was to test whether infant gonadotropin concentrations were associated with later spontaneous puberty as well as determine the threshold of infant LH and FSH that best predicts the occurrence of spontaneous menarche and thelarche. LH and FSH are not normally distributed in our cohort, therefore medians with interquartile ranges (IQR) are reported and differences in infant values were determined between patients who did and did not undergo spontaneous thelarche or menarche using Kruskal-Wallis tests. The optimal LH and FSH thresholds for predicting spontaneous thelarche and menarche were determined as the value that maximized the sum of sensitivity and specificity. A bootstrap determination of the threshold was calculated using the cutpointr package in R.(14) From the optimal threshold, we report the associated bootstrap estimated sensitivity, specificity, and accuracy. The aim of the secondary analysis was to compare infant LH and FSH to childhood AMH concentrations. Given the floor effect of AMH, Kruskal-Wallis tests were used to test if infant LH and FSH differed in patients with an undetectable AMH compared to patients with a quantifiable AMH. A type 1 error rate of 0.05 is assumed. All statistical analyses were performed in R, version 4.3.2.

## Results

A total of 33 patients met inclusion criteria with a mean age of 16.2 years at time of data collection (Table 1). Karyotype 45,X represents the largest proportion of included patients with 48.5% while 15.2% had a karyotype of 45,X/46,XX, and 36.2% had a variety of other karyotypes. Spontaneous thelarche occurred in 21.2% and spontaneous menarche occurred in 15.2% of the cohort.

**Table 1.** Cohort Demographics.

|  | Overall<br>(N=33) |
| --- | --- |
| **Age at Last Visit (Years)** |  |
| Mean (SD) | 16.2 (3.8) |
| **Insurance** |  |
| Private/Commercial | 19 (57.6%) |
| Medicaid | 14 (42.4%) |
| **Race** |  |
| Asian | 2 (6.1%) |
| Black/African American | 1 (3.0%) |
| White | 26 (78.8%) |
| More than One Race | 4 (12.1%) |
| **Ethnicity** |  |
| Hispanic or Latino(a) | 9 (27.3%) |
| Not Hispanic or Latino(a) | 24 (72.7%) |
| **Spontaneous Thelarche** |  |
| Yes | 7 (21.2%) |
| No | 26 (78.8%) |
| **Spontaneous Menarche** |  |
| Yes | 5 (15.2%) |
| No | 23 (69.7%) |
| Age not applicable<sup>1</sup> | 5 (15.2%) |
| **Karyotype** |  |
| 45,X | 16 (48.5%) |
| 45,X/46,XX | 5 (15.2%) |
| 45,X/46,XY | 1 (3.0%) |
| 45,X/47,XXX | 2 (6.1%) |
| Ring X | 1 (3.0%) |
| Isochromosome X | 4 (12.1%) |
| Autosomal Translocation | 1 (3.0%) |
| Deletion on the X chromosome | 3 (9.1%) |
| **Timing of TS Diagnosis** |  |
| Prenatal | 14 (42.4%) |
| Postnatal | 19 (57.6%) |

<sup>1</sup>Not exposed to exogenous estrogen and not yet clinically abnormal to not have reached menarche ( $\leq 15$ years of age and normal gonadotropins)

Infant LH strongly correlated with infant FSH, and both were predictive of later ovarian function outcomes (Figure 1). Patients who underwent spontaneous thelarche had significantly lower infant FSH values (12.8 mIU/mL [5.0, 27.7] vs 79.2 mIU/mL [57.9, 111.9], p=0.001) and LH values (0.5 mIU/mL [0.2, 1.7] vs 4.3 mIU/mL [2.6, 8.0], p=0.004) compared to patients who did not undergo spontaneous thelarche (Figure 2). Patients who underwent spontaneous menarche also had significantly lower infant FSH values (12.8 mIU/mL [7.9, 17.9] vs 82.6 mIU/mL [59.3, 109.8], p<0.001) and LH values (0.5 mIU/mL [0.2, 0.5] vs 4.0 mIU/mL [2.7, 8.1], p=0.008) compared to patients who did not undergo spontaneous menarche. The significant associations between infant LH and FSH and later ovarian function outcomes remained after adjusting for mosaicism status. Childhood AMH values were available for 24 of the 33 patients. While the earliest available childhood AMH values ranged from undetectable to 3.3 ng/ml, 70.8% were undetectable. Patients with an undetectable childhood AMH had a significantly higher infant FSH (82.6 mIU/mL [59.4, 87.2] vs 17.9 mIU/mL [10.4, 51.4], p=0.008) and LH (3.3 mIU/mL [2.8, 7.0] vs 0.5 mIU/mL [0.19, 3.4], p=0.049).

**Fig 1.**
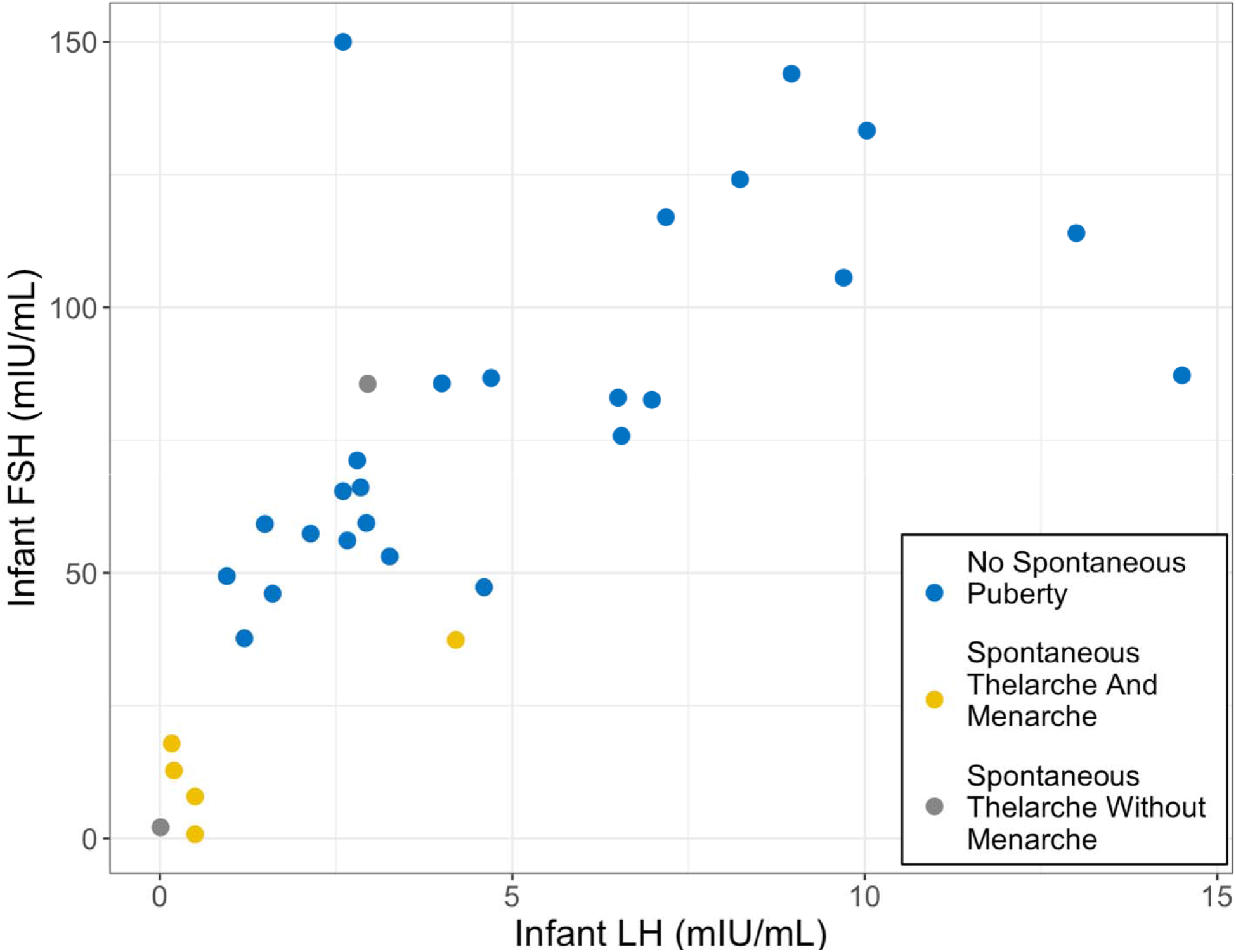
Scatterplot of infant FSH by LH stratified by pubertal status. Each dot represents an individual patient. LH values range from 0.01 to 14.5 mIU/mL. One additional person had an infant LH of 49.9 and an FSH of 170.

**Fig 2.**
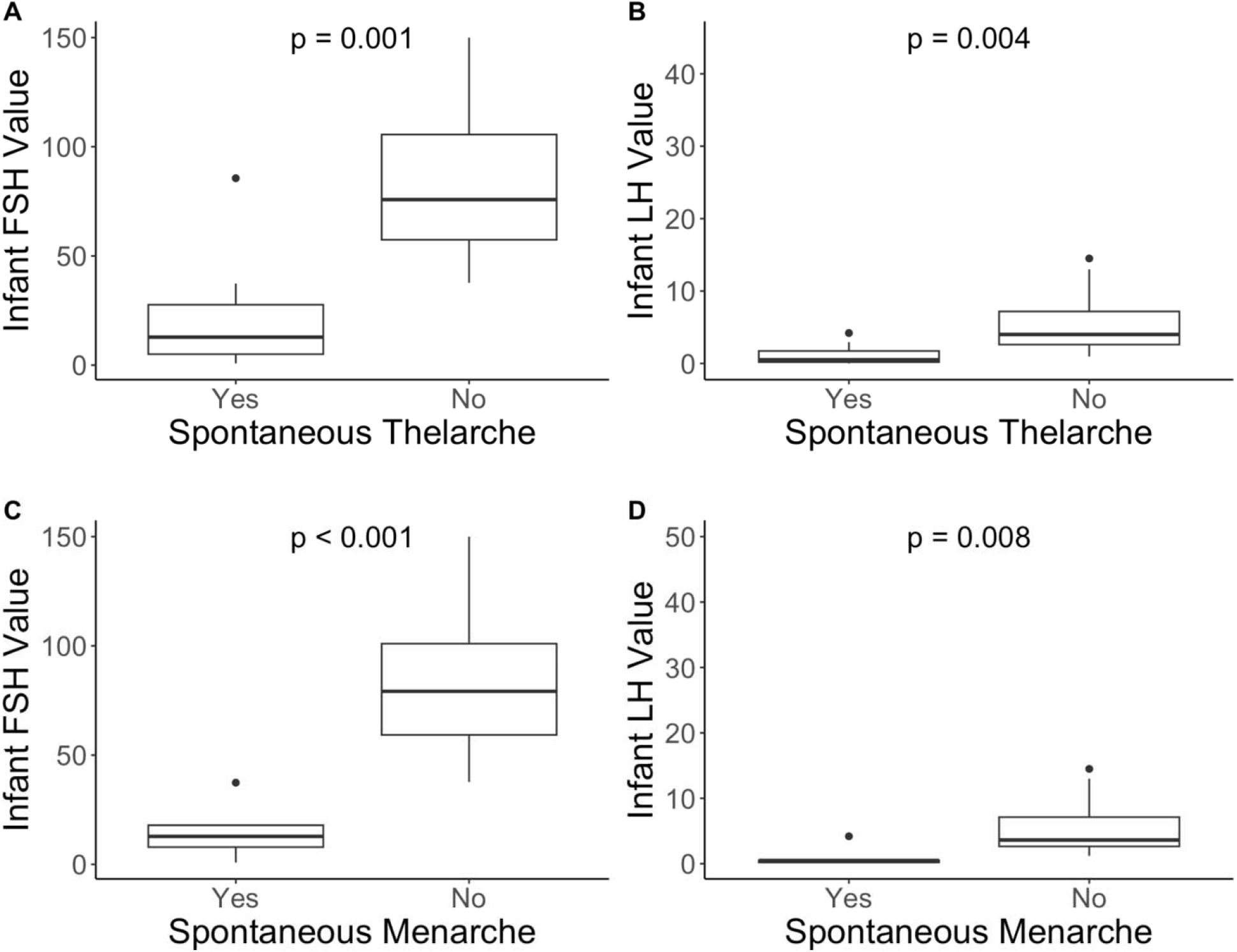
LH and FSH concentrations for patients that underwent spontaneous thelarche (A, B) and spontaneous menarche (C, D) compared to patients that did not undergo spontaneous menarche or thelarche. P-values represent Kruskal-Wallis tests for the difference in FSH/LH values between patients who did and did not undergo spontaneous menarche/thelarche. One additional person had an infant LH of 49.9. Significance level = 0.05.

**Fig 3.**
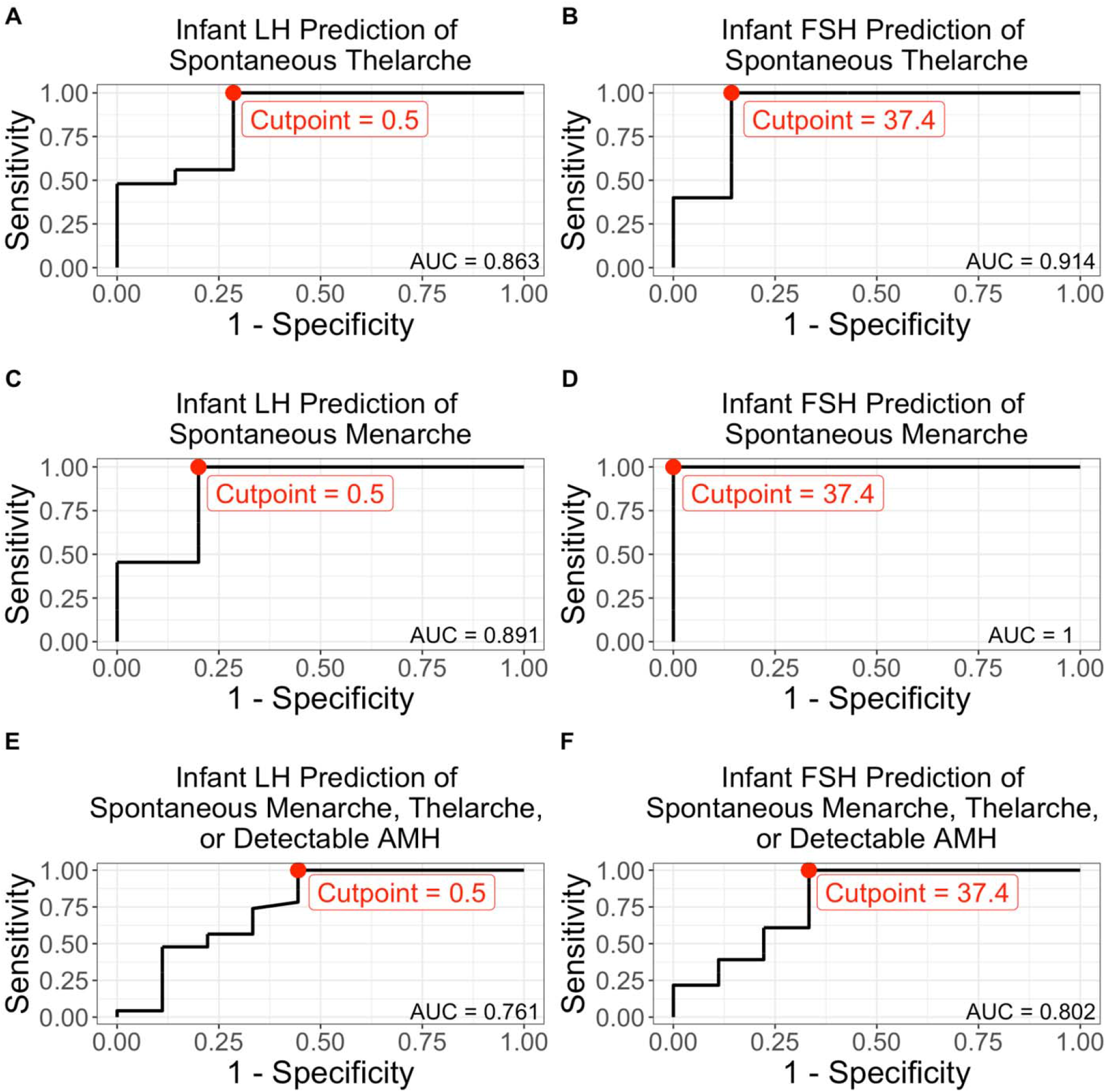
Receiver operating characteristic (ROC) curves where red points indicate the cut point that maximizes the sum of sensitivity and specificity. a) infant LH (<0.5 mIU/mL) as a predictor of spontaneous menarche (sensitivity 0.8 specificity 1.0, accuracy 96.4%) b) infant FSH (<37.4 mIU/mL) as a predictor of spontaneous menarche (sensitivity 1.0, specificity 1.0, accuracy 100%) c) infant LH (<0.5 mIU/mL) as a predictor of spontaneous thelarche (sensitivity 0.71 specificity 1.0, accuracy 93.9%) d) infant FSH (<37.4 mIU/mL) as a predictor of spontaneous thelarche (sensitivity 0.86, specificity 1.0, accuracy 97.0%) e) infant LH (<0.5 MIU/L) as a predictor of spontaneous menarche, thelarche, or detectable AMH (sensitivity 0.56 specificity 1.0, accuracy 87.5%) f) infant FSH (<37.4 mIU/mL) as a predictor of spontaneous menarche, thelarche, or detectable AMH (sensitivity 0.67 specificity 1.0, accuracy 90.6%)

An infant LH cut-off value of <0.5 mIU/mL was found to predict spontaneous thelarche most optimally (N = 33) with a sensitivity of 71.4% (Bootstrap 95% CI: 57%, 100%), specificity of 100% (Bootstrap 95% CI: 54%, 100%), and overall accuracy of 93.9% (Bootstrap 95% CI: 64%, 100%). The same infant LH cut-off value of <0.5 mIU/mL was found to predict spontaneous menarche most optimally (N = 28) with a sensitivity of 80% (Bootstrap 95% CI: 50%, 100%), specificity of 100.0% (Bootstrap 95% CI: 62%, 100%), and overall accuracy of 96.4% (Bootstrap 95% CI: 68%, 100%). Similarly, to predict spontaneous menarche, thelarche, or detectable AMH in childhood or puberty, an infant LH cut-off value of <0.5 mIU/mL was most optimal (N = 33) with a sensitivity of 55.6% (Bootstrap 95% CI: 17%, 91%), specificity of 100% (95% CI : 54%, 100%), and overall accuracy of 87.5% (Bootstrap 95% CI : 62%, 97%).

An infant FSH cut-off value of <37.4 mIU/mL was found to predict spontaneous menarche most optimally (N=28) with a sensitivity of 100% (Bootstrap 95% CI: 100%, 100%), specificity of 100% (Bootstrap 95% CI: 100%, 100%), and overall accuracy of 100% (Bootstrap 95% CI: 100%, 100%). The same infant FSH cut-off value of <37.4 mIU/mL was found to predict spontaneous thelarche most optimally (N=33) with a sensitivity of 85.7% (Bootstrap 95% CI: 60%, 100%), specificity of 100% (Bootstrap 95% CI: 100%, 100%), and overall accuracy of 97.0% (Bootstrap 95% CI: 91%, 100%). Similarly, to predict spontaneous menarche, thelarche, or detectable AMH in childhood or puberty, an infant FSH cut-off value of <37.4 mIU/mL was most optimal (N = 33) with a sensitivity of 66.7% (Bootstrap 95% CI: 43%, 100%), specificity of 100% (95% CI : 82%, 100%), and overall accuracy of 90.6% (Bootstrap 95% CI : 79%, 100%).

## Discussion

This retrospective longitudinal cohort analysis demonstrated that infant FSH and LH can accurately predict later ovarian function in patients with TS. Specifically, an infant FSH cut off level of 37.4 mIU/mL and an LH of 0.5 mIU/mL predicted spontaneous thelarche and menarche in adolescents with TS with a high sensitivity, specificity and overall accuracy. We also confirmed a significant relationship between infant FSH and LH and later childhood AMH. This is the first study, to our knowledge, to find that infant gonadotropin measurements may have utility for predicting later ovarian function outcomes in girls with TS.

A previous systematic review found rates of spontaneous thelarche to be 32% and spontaneous menarche to be 21% in girls with TS.(5) We saw slightly lower rates in our study population (21.2% and 15.2% respectively), likely because of the high proportion of our sample that had non-mosaic 45,X. Our cohort was born an average of 16 years ago, prior to the adoption of non-invasive prenatal screening into clinical care, therefore most girls who had a diagnosis of TS prior to 3 years of age (and subsequently had gonadotropins measured and included in this study), were diagnosed early due to a more severe phenotypic TS presentation. Karyotype is a known predictor for spontaneous puberty in TS; mosaicism with 46,XX doubles the chance of spontaneous puberty over non-mosaic 45,X.(4,5) While karyotype can provide insight into the chance of spontaneous puberty, there is still a great deal of individual variability that limits ability to confidently counsel parents. Additional biomarkers, such as infant gonadotropins, can be helpful for more individualized counseling.

Prior studies have found a pattern of elevated gonadotropins in early childhood for girls with TS, followed by a nadir in mid-childhood and another rise in adolescence.(15) However, there is a significant overlap with normal levels. Although logical, it has not previously been clear if elevated infant gonadotropins accurately reflect irreversible primary ovarian failure and therefore absence of future puberty and fertility. Our study provides these missing data so that clinicians can use gonadotropins obtained in infancy to provide more individualized information to parents earlier in life. Indeed, elevated gonadotropins in infancy are predictive of POI as assessed by absence of spontaneous puberty in the TS population.

As a secondary finding, we demonstrated infant gonadotropins also predict later childhood AMH. AMH is a growth factor made by the granulosa cells in developing primordial follicles in the ovary that correlates with the size of the follicular pool and thus can reflect ovarian reserve and aid in fertility counseling.(9) In adolescents and adults with TS, an AMH <0.42 ng/mL has been found to be a specific and sensitive marker of POI.(16,17) Furthermore, in the TurnerFertility trial, a detectable childhood AMH had the strongest correlation with the presence of follicles in ovarian tissue compared to the other biomarkers, and AMH correlated with the number of follicles.(13) Therefore, AMH is likely a helpful marker to potentially inform which patients may be the best candidates for ovarian tissue cryopreservation (OTC). Our observation that infant gonadotropins can predict detectable childhood AMH may be helpful information for early discussions on fertility. OTC is still an experimental procedure in TS and more research is needed to understand the chance of pregnancy from OTC in this population.(16) Most OTC research protocols require girls with TS to be >2 years of age, however, elevated infant gonadotropins may be an indication to consider OTC for infants already undergoing anesthesia cardiac, otolaryngology, or other surgical procedures, which are common in this population.(1) Whether or not AMH in infancy could serve the same role as infant gonadotropins will need to be explored, as our cohort is too old to have had AMH measured on a clinical basis while they were infants.

A major strength of this study includes the longitudinal data collection from infancy through at least 10 years of age. As this was a clinical convenience sample there is overrepresentation of individuals with an early TS diagnosis, which likely biases toward those with greater phenotypic impact given the timing was prior to implementation of routine cell-free DNA screening. Gonadotropin and AMH measures were obtained clinically at more than one commercial laboratory, therefore there is inevitable heterogeneity that may limit generalizability of the specific cut-off value. The small sample size and limited duration of follow up restricts our outcome to spontaneous puberty only, and we cannot comment on ovarian function after spontaneous pubertal onset. Further studies are needed to assess the natural history of ovarian function throughout the lifespan in TS.

## Conclusions

In conclusion, we have found infant FSH and LH to be accurate biomarkers of future ovarian function in individuals with TS, as defined by spontaneous puberty and childhood AMH concentrations. This study supports measuring gonadotropins within the first years of life to provide more individualized counseling and anticipatory guidance on puberty and fertility for the TS population. In addition, clinical interventions targeting fertility preservation should consider infant gonadotropins as reliable biomarker for later ovarian function.

## Data Availability

Data are available from the authors upon reasonable request.

## Acknowledgements

We want to acknowledge the non-profit organization Turner Syndrome Colorado that supports the eXtraOrdinary Kids Turner Syndrome Multidisciplinary Clinic at Children’s Hospital Colorado and all the patients and providers that are a part of that clinic.

